# Interaction-based feature selection algorithm outperforms polygenic risk score in predicting Parkinson’s Disease status

**DOI:** 10.1101/2021.07.20.21260848

**Authors:** Jörn E. Klinger, Charles N. J. Ravarani, Hannes A. Baukmann, Justin L. Cope, Erwin P. Böttinger, Stefan Konigorski, Marco F. Schmidt

## Abstract

Polygenic risk scores (PRS) aggregating results from genome-wide association studies are state of the art to predict the susceptibility to complex traits or diseases. Novel machine learning algorithms that use large amounts of data promise to find gene-gene interactions in order to build models with better predictive performance than PRS. Here, we present a data preprocessing step by using data-mining of contextual information to reduce the number of features, enabling machine learning algorithms to identify gene-gene interactions. We applied our approach to the Parkinson’s Progression Markers Initiative (PPMI) dataset, an observational clinical study of 471 genotyped subjects (368 cases and 152 controls). With an AUC of 0.85 (95% CI = [0.72; 0.96]), the interaction-based prediction model outperforms the PRS (AUC of 0.58 (95% CI = [0.42; 0.81])). Furthermore, feature importance analysis of the model provided insights into the mechanism of Parkinson’s Disease. For instance, the model revealed an interaction of previously described drug target candidate genes *TMEM175* and *GAPDHP25*. These results demonstrate that interaction-based machine learning models can improve genetic prediction models and might provide an answer to the missing heritability problem.

## Introduction

The need to understand how to predict phenotypes from genetic data becomes ever-more important for individual’s disease risk prediction, animal and plant breeding as well as genome editing. Polygenic risk scores (PRS), simple additive models, are state of the art to investigate the genetic architecture and, more importantly, to predict the susceptibility of complex traits or diseases. (Wray et al., 2007; Evans et al., 2009; International Schizophrenia Consortium et al., 2009) For each individual a score is calculated as a weighted sum of the number of risk allele single nucleotide polymorphisms (SNP) an individual was tested for. The used weights are regression coefficients from previous genome-wide association studies (GWAS).

Importantly, PRS models are not optimized for predictive performance. (Chatterjee et al., 2013; Dudbridge, 2013) There are three reasons for this:

1. Due to the current limited sample size of discovery GWAS datasets (< 1,000,000 individuals), biologically relevant rare variants with small effect sizes cannot be detected. Additionally, the limited sample sizes of discovery GWAS can lead to biased PRS models that might not perform well in populations with ancestry different to that of the discovery dataset. (Reisberg et al., 2017; Duncan et al., 2019)
2. It has been shown that statistically associated SNPs are not automatically good predictors. (Lo et al., 2015)
3. It has been reported that genetic effects discovered in genome-wide association studies do not sum to the estimate of the heritability of the trait compared to twin studies. (Yang et al., 2010) This has been called the missing heritability problem in GWAS. (Manolio et al., 2009) Beside potentially missing relevant rare variants and suboptimal SNP selection based on *p* values, classical PRS models ignore complex gene-gene interactions, also known as epistasis, of the trait or disease due to their simple additive structure.

The concept of epistasis has been described more than 100 years ago. (Bateson and William, 1906) Statistical epistasis, as observed in genome-wide association studies, is genetic variance that can be attributed to gene interaction and is defined as a function of the allele frequencies in a population. Detection of epistasis in discovery GWAS and modeling its impact is challenging because of linkage disequilibrium (LD), replication of identified gene-gene interactions in validation datasets, model complexity, and high dimensionality. (Wei et al., 2014)

Machine learning algorithms that improve automatically through experience and by the use of data represent an opportunity to find gene-gene interactions in order to build prediction models with better predictive performance than PRS. Nevertheless, in a recent study a PRS model outperformed the five machine learning algorithms Naïve Bayes classifier, regularized regression, random forest, gradient boost, and support vector machine used to build prediction models for coronary artery diseases status. (Gola et al., 2020)

Here, we explore the potential of using contextual information obtained via data mining to strongly reduce the hypothesis space, which, in turn, allows for testing a small set of complex hypotheses, containing interaction of multiple variants. This approach organizes data mined from journal articles, pathway libraries, protein co-expression libraries and drug candidate libraries into a hierarchical graph, which generates disease-specific hypotheses based on interactions of genetic variants. Each interaction’s predictive power is determined using the training data set. If an interaction predicts disease status well, the graph is incentivized to ‘fine-tune’ the hypothesis by comparing a set of very similar hypotheses. If a hypothesis has little or no predictive power, the graph is not incentivized to explore it, or similar hypotheses further and will instead propose hypotheses containing different variants. This learning process is driven by gradient descent, meaning that it converges when the average performance of the new multi-variant hypothesis does not increase. After convergence, the selected features are used to build prediction models with standard machine learning algorithms, such as LASSO (least absolute shrinkage and selection operator) regression (Friedman et al., 2010). An overview of our approach is given in Figure 1. The Parkinson’s Progression Markers Initiative (PPMI) dataset (Marek et al., 2011, 2018) (https://www.ppmi-info.org) was selected for the comparison as the dataset has been intensively analyzed and, from there, a broad audience is able to reproduce our results.

**Fig. 1.**
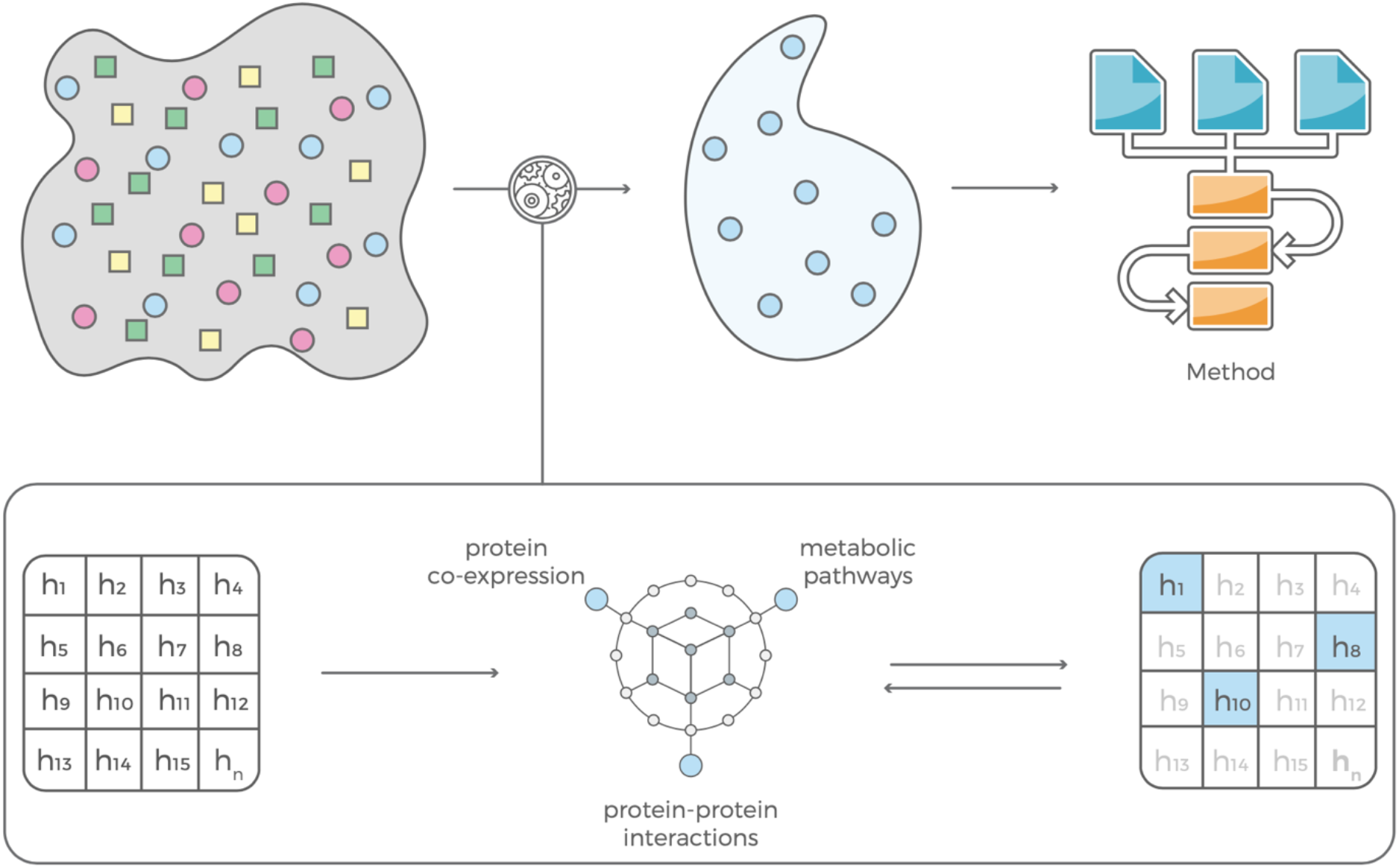
Our feature selection consists of two complementary modules that are in feedback with each other. The contextual module uses information mined from the scientific literature, pathway libraries and protein co-expression data and an evaluation module that estimates predictive power of a feature based on that contextual information. The selected features can be used to build prediction models with standard machine learning algorithms.

## Results

### GWAS

In a preliminary proof-of-concept step, a genome-wide association (GWA) analysis was performed. For all 471 subjects in the PPMI database, 368 cases and 152 controls, subject genotyping information was collected from two complementary genotyping chips (NeuroX and ImmunoChip). After careful quality control and harmonization, we merged that information into a single dataset with 369,036 variants and 436 individuals. The Manhattan plot of the *p* values resulting from SAIGE analysis is shown in Fig. 2. Seven single nucleotide polymorphisms (SNPs) showed smaller *p* values than 10^−4^ (Table 1).

**Tab 1.** PPMI GWAS results identified 7 SNPs with a p value < 10^−4^. Positions and rs IDs according to Human Genome Reference hg19 (GRCh37).

| Ch<br>r | Pos | SNP ID | rs ID | Genic Context | $p$ value |
| --- | --- | --- | --- | --- | --- |
| 1 | 173266578 | imm_1_17<br>1533201 | rs4916319 | *TNFSF4* (upstream) | 0.000083 |
| 2 | 209087335 | exm22611<br>59 | rs4675743 |  | 0.000046 |
| 5 | 156376703 | exm49891<br>7 | rs6873053 | *TIMD4* (downstream) | 0.000092 |
| 6 | 133716974 | rs212805 | rs212805 | *EYA4* | 0.000074 |
| 17 | 25895033 | imm_17_2<br>2919160 | rs4795747 |  | 0.000015 |
| 18 | 5479093 | rs7238186 | rs7238186 | *EPB41L3* (downstream) | 0.000007 |
| 19 | 57909872 | exm15132<br>84 | rs4801478 | *ZNF548* | 0.000040 |

**Fig. 2.**
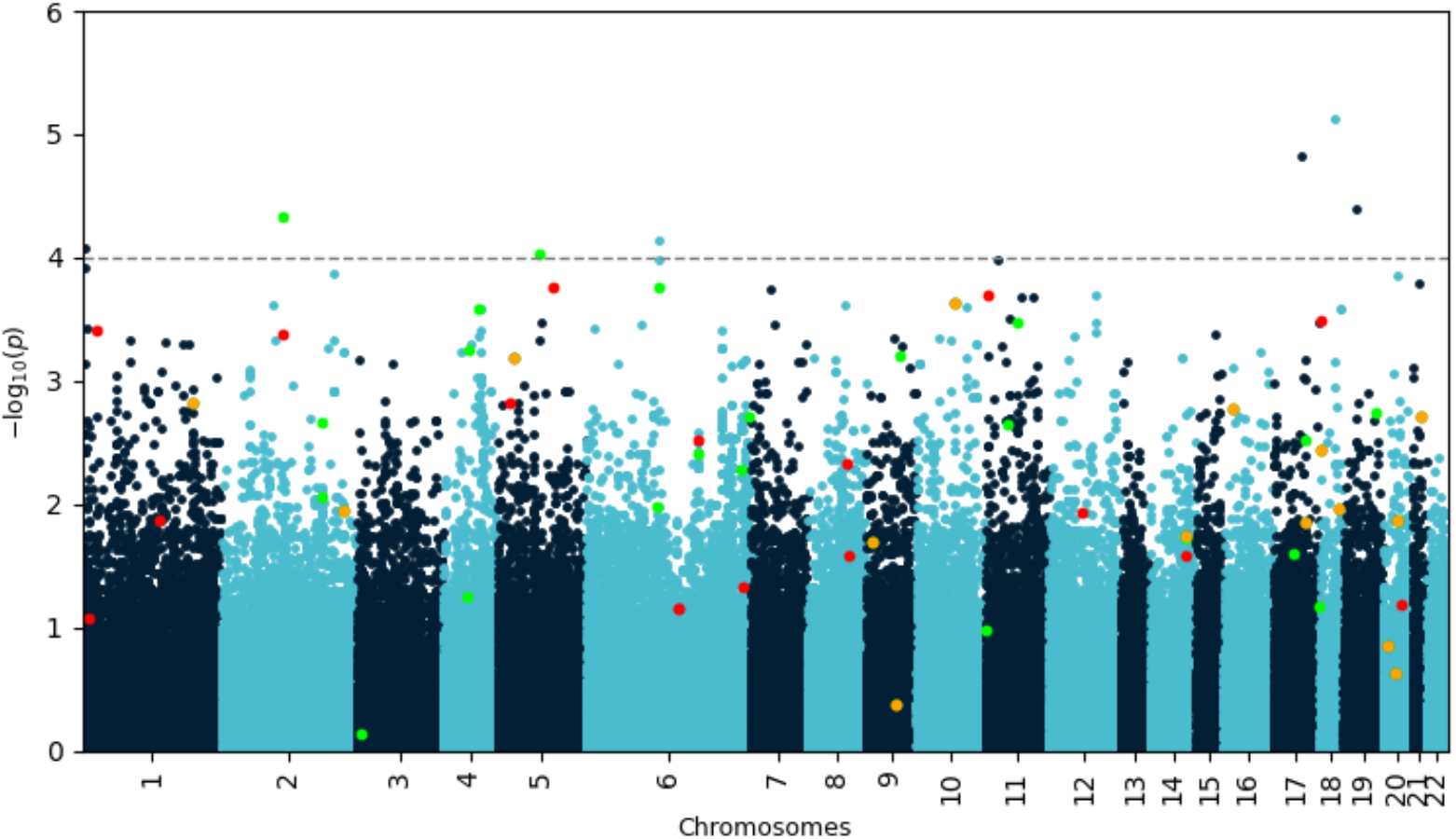
Manhattan plot of negative decadic logarithm of *p* values for SNPs as determined by SAIGE analysis. Variants identified by our biotx.ai model are highlighted in red and green if they increase or decrease disease risk, respectively. Variants highlighted in orange occur in both protective and risk-enhancing groups of SNPs, depending on their genotype. Most of these biologically meaningful variants would have been missed by using a simple *p* value cutoff.

**Fig. 3.**
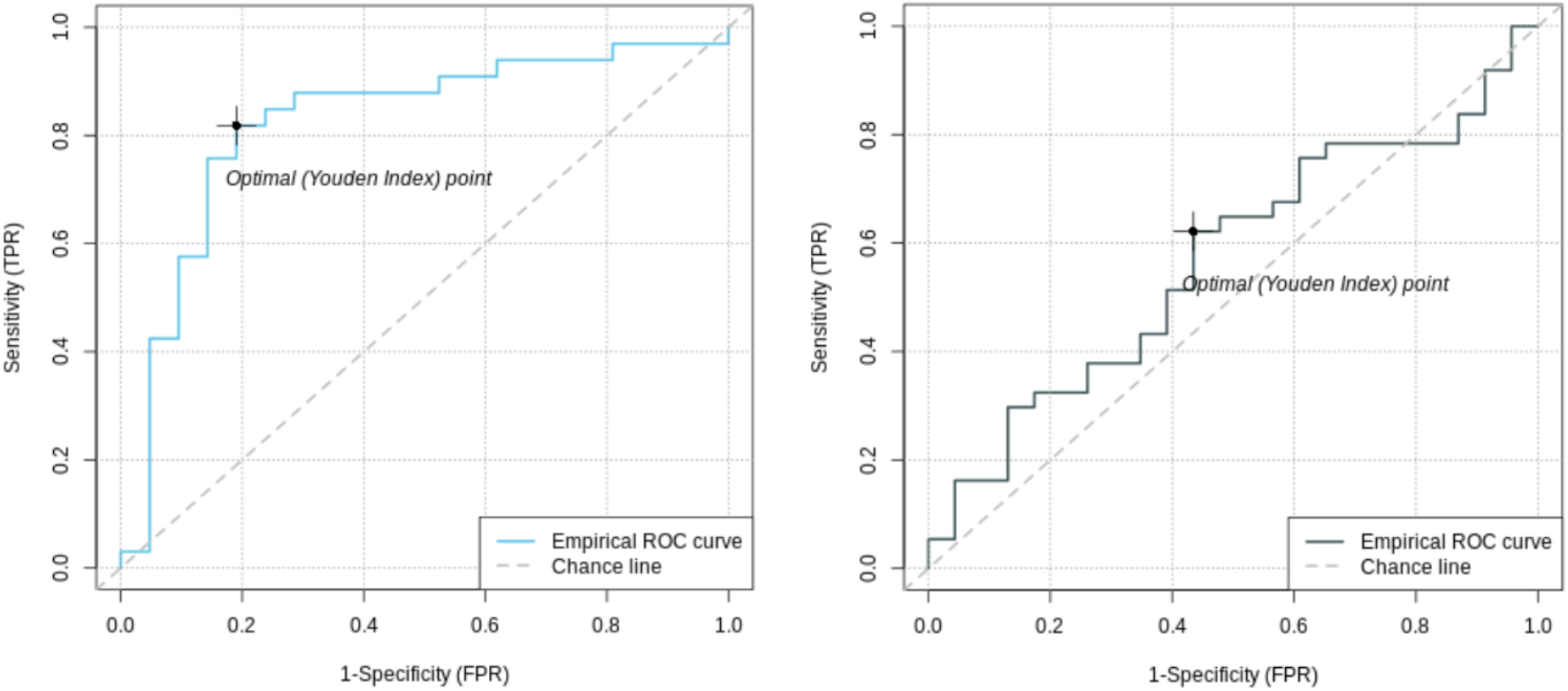
Receiver operating characteristic (ROC) curves of feature selected machine learning model (left) and polygenic risk score (right). The AUC of the feature selected model with 0.85 (95% CI = [0.72; 0.96]) is better than the AUC of the PRS with 0.56 (95% CI = [0.42; 0.81]).

**Fig. 4.**
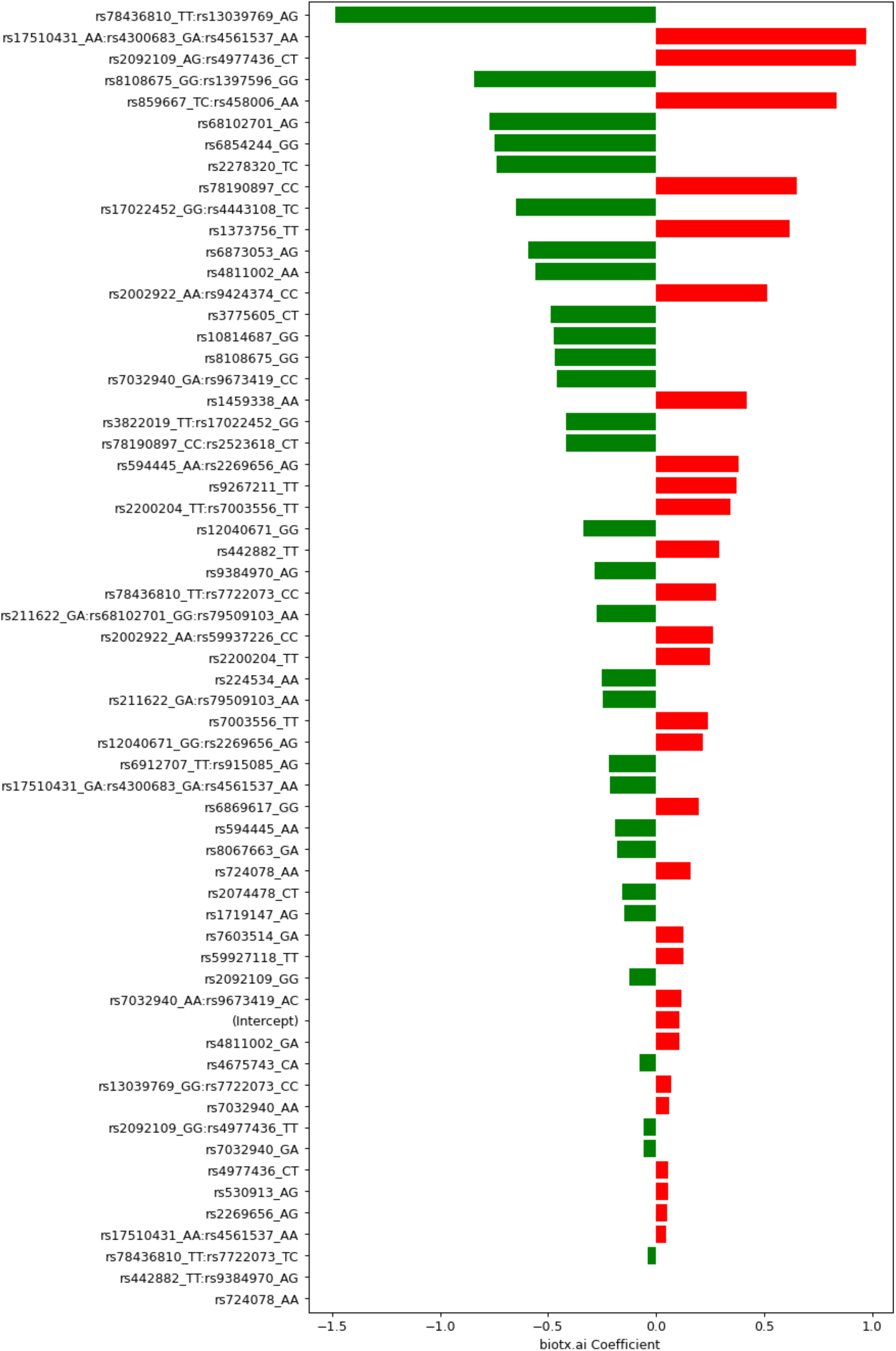
Coefficients determined by our biotx.ai model for SNPs and groups of SNPs. Negative values (green) indicate protective (combinations of) variants, positive values (red) mark risk variants. The respective genotypes of each variant are indicated by one-letter codes of the bases, where the first letter corresponds to the reference allele, and the second corresponds to the observed, alternative allele.

### Polygenic risk score

For all analyses described in the following, the data was split into training, validation and test sets. The same sets were used for constructing a polygenic risk score (PRS) and machine learning prediction models with and without feature selection. To calculate the PRS, 7 different *p* value thresholds (0.001, 0.05, 0.1, 0.2, 0.4, 0.5) for the subjects in the training, validation and test set were used. The PRS of the subjects in the training set were then used to train a separate logistic regression classifier for each *p* value threshold. The validation data set was used to determine which of these thresholds produces the best classifier, which was then used to predict the test set. This classifier is based on the PRS of 57 different SNPs.

The receiver operating characteristics (ROC) curve was used to evaluate the predictive power of the PRS. The area under the curve (AUC) was 0.58 (95% CI = [0.42; 0.81]) and the Youden’s index was 0.21 (Tab. 2).

### Deep learning

Deep learning is a machine learning technique based on artificial neural networks with representation learning that allows a system to automatically discover the representations needed for feature detection or classification from raw data. Despite not being widely used in the field of genomics, there is work on applying Deep Learning to GWAS, such as Romero *et al. (Romero et al., 2016)*. Romero *et al*. use a Diet Network, a neural network parameterization, which considerably reduces the number of free parameters. The model is composed of 3 networks, one basic and two auxiliary networks. After a basic discriminative network with optional reconstruction path, a network that predicts the input fat layer parameters, and finally, a network that predicts the reconstruction fat layer parameters. We applied their approach to the PPMI dataset. The area under the curve (AUC) of the deep learning model was 0.67 (95% CI = [0.47; 0.83]) and the Youden index was 0.29 (Tab. 2). Notably, the deep learning model contains abstract embeddings instead of concrete SNPs as in the PRS.

**Tab. 2.** Performance comparison of all models.

| Method | AUC<br>[95% CI] | Accuracy | Sensitivity | Specificity | Youden'<br>s Index |
| --- | --- | --- | --- | --- | --- |
| PRS | 0.56<br>[0.42; 0.81] | 0.60 | 0.62 | 0.56 | 0.21 |
| Deep learning | 0.67<br>[0.47; 0.83] | 0.60 | 0.42 | 0.88 | 0.29 |
| LASSO w/<br>feature selection | 0.85<br>[0.72; 0.96] | 0.81 | 0.81 | 0.80 | 0.61 |
| LASSO w/o<br>feature selection | 0.51<br>[0.39; 0.63] | 0.62 | 0.87 | 0.09 | 0.12 |

### Feature selection and LASSO regression

The hierarchical graph, as well as the training set of the GWAS data were used to select a set of less than 100 polygenic hypotheses using our approach as described in the introduction above. The remaining hypotheses were summarized in a term that was used to train a LASSO regression model on the validation data. (Tibshirani, 1996) This model, based on 47 SNPs in several different interaction terms, then predicted the test set. The area under the curve (AUC) for the LASSO model with prior feature selection was 0.85 (95% CI = [0.72; 0.96]) and the Youden index was 0.61. A LASSO model without prior feature selection that was built for comparison did not deliver outcomes that were significantly better than chance (Tab. 2).

Exploring the feature selection based model with its interactive terms, provides insights about the genes associated with the disease. An annotation of all 47 SNPs in our model can be found in the Supplementary Information. An exciting result from this analysis of the PPMI dataset is the statistical interaction of variants rs3822019 on chromosome 4 in gene *TMEM175*, coding for a potassium channel in late endosomes, and rs17022452 on chromosome 2, close to the coding region of *GAPDHP25*, glyceraldehyde-3 phosphate dehydrogenase pseudogene 25. rs3822019 is an intron variant that has been linked to Parkinson’s Disease. (Nalls et al., 2014)

## Discussion

We analyzed the PPMI dataset and built predictive models using PLINK for a polygenic risk score, a diet-net deep learning algorithm for genomic data (Romero et al., 2016), and LASSO regression with and without the above proposed approach of using contextual data to reduce the hypothesis space. The PRS model comprises 16,135 SNPs and showed an AUC of 0.56 whereas the deep learning model had an AUC of 0.52. Notably, the deep learning model consists of abstract embeddings instead of single SNPs like the PRS. Therefore, identification of disease-associated SNPs and further insights into the disease mechanism are not possible here. The LASSO regression model built on interactions containing only 47 SNPs that were discovered via the use of contextual information outperformed the other predictive models with an AUC of 0.82. Beyond that, the approach was able to associate new variants with the disease that would have not shown up under an additive approach such as PRS. We investigated how the combinations of the relevant genotypes rs3822019_TT (*TMEM175*) and rs17022452_GG (*GAPDHP25*) split the individuals into cases and controls (Tab. 3). All subjects that are homozygous for rs3822019_TT are affected by PD. Furthermore, most individuals heterozygous for this variant (rs3822019_TT) or homozygous for rs17022452_GG are cases (76.4% and 75.0%, respectively). These results support the relevance of the association between these variants and PD status. The *TMEM175/GAK/DGKQ* locus was the third strongest risk locus in a GWA study of Parkinson’s disease (Krohn et al., 2020) and has been described as a potential drug target. (Diogo et al., 2018; Jinn et al., 2019) Deficiency in the potassium channel TMEM175 results in unstable lysosomal pH, which leads to decreased lysosomal catalytic activity and increased α-synuclein aggregation, among other effects. As a potassium channel, TMEM175 has a high potential as a druggable target anda tractable therapeutic strategy has been proposed. (Jinn et al., 2017)

**Tab. 3.** PD cases and controls among bearers of the respective genotype combinations of the identified variants rs3822019 and rs17022452.

| Genotype combination | cases | controls |
| --- | --- | --- |
| rs3822019_TT / rs17022452_GG | 0 | 0 |
| rs3822019_TT / rs17022452_GA | 6 / 100% | 0 |
| rs3822019_TC / rs17022452_GG | 2 / 50% | 2 / 50% |
| rs3822019_TT / - | 7 / 100% | 0 |
| - / rs17022452_GG | 7 / 87.5% | 1 / 12.5% |
| rs3822019_TC / rs17022452_GA | 27 / 87.1% | 4 / 12.9% |
| rs3822019_TC / - | 68 / 73.9% | 24 / 26.1% |
| - / rs17022452_GA | 66 / 75% | 22 / 25% |
| - / - | 113 / 56.5% | 87 / 43.5% |

GAPDH has been targeted with the investigational drug Omigapil for prevention of PD, ALS, congenital muscular dystrophy and myopathy. The drug has been shown to protect against behavioural abnormalities and neuro-degeneration in animal models of Parkinson’s disease. However, PD development has been terminated due to lack of benefit. (Olanow et al., 2006)

There seem to be various causes of Parkinson’s Disease, yet the pathogenesis of this disease appears to be converging on common themes—oxidative stress, mitochondrial dysfunction, and protein aggregation—all of which are tightly linked to autophagy. (Lynch-Day et al., 2012) Both TMEM175 (Jinn et al., 2019) and GAPDH(Butera et al., 2019) regulate autophagy. Disturbed expression of autophagy genes in blood of PD patients. (Lynch-Day et al., 2012)

To summarize, we here present an approach to apply machine learning algorithms to high-dimensional genomic data using a contextual knowledge based feature selection. PRS models require a large set of SNPs, which leads to overfitting and limits their use in clinical practice. We generated more parsimonious models overcoming these limitations – with only 47, partly interacting SNPs, our model was able to outperform a PRS model based on 57 SNPs for Parkinson’s Disease. Analysis of feature importance of our model identified a gene-gene interaction of *TMEM175* and *GAPDHP25*. TMEM175 has been described as a potential drug target and further information on its mechanism of action could be invaluable. A recently discovered interaction with pseudogene *GAPDHP25* could provide helpful insights. In conclusion, applying machine learning algorithms to feature-selected genomic data leads to interaction-based models with better predictive performance than PRS as well as paves the way to generate new insights into disease mechanisms.

## Methods

### Parkinson’s progression marker initiative dataset

The Parkinson’s progression marker initiative (PPMI) dataset (https://www.ppmi-info.org) contains 471 subjects, 368 cases and 152 controls, for each subject genotyping information from two complementary chips (NeuroX and ImmunoChip) was collected. (Marek et al., 2011) After careful quality control and harmonization (e.g. genome build, strand alignment) as described in the literature (Marees et al., 2018) we merged that information into a single dataset with 380,939 variants in total. An additional set of quality control steps were performed on variants and individuals that aimed to remove biases that could affect the downstream analysis.

In more detail, in a first stage SNPs and individuals were filtered based on their missingness in the dataset. This ensures that SNPs are excluded that have a high proportion of subjects where genotyping information was unavailable or of poor quality. Similarly, individuals where a large proportion of SNPs could not be measured were excluded. This step was achieved by setting a threshold of 0.02 (i.e. >2%; 6,084 variants and 22 people were removed). SNP filtering was performed before individuals were filtered.

With high missing rates filtered, all variants not on autosomal chromosomes were removed (5,731 variants were removed). This was followed by the identification and removal of variants that deviate from the Hardy-Weinberg equilibrium. These variants were identified in a two-stage process whereby we first applied a threshold of 1e-6 exclusively to controls, followed by a threshold of 1e-10 applied to all samples (0 and 202 variants were removed). This is a common indicator of genotyping errors.

Next, individuals were filtered based on their heterozygosity rates which can indicate sample contamination. Individuals that deviate by more than 3 standard deviations from the mean of the rate from all samples (13 individuals were removed) were filtered out. To assess the heterozygosity rate per sample, the variants that were in linkage disequilibrium with each other were first extracted, scanning the genome at a window size of 50 variants, step size of 5 and a pairwise correlation threshold of 0.2.

Finally, related individuals were removed, which was achieved through the assessment of their respective identity by descent coefficients (IBD) that were calculated. Only one individual in a related pair would be kept (0 individuals were removed).

The final quality-controlled dataset contained 369,036 variants and 436 individuals passing the various filters.

### GWAS

In a preliminary proof-of-concept step, a genome-wide association (GWA) analysis was performed with the R package SAIGE (Zhou et al., 2018) to test individual variants for their association with Parkinson’s Disease.

### Polygenic risk score

The data (n = 436) was then split into training (n = 367), validation (n = 33) and test sets (n = 36). The same sets were used for constructing a polygenic risk score (PRS) and machine learning prediction models with and without feature selection. To calculate the PRS, different *p* value thresholds for the subjects in the training, validation and test set were used. The PRS was constructed by using PLINK(Purcell et al., 2007) following the guidelines provided by Choi et al. (Choi et al., 2020) and the accompanying tutorial (https://choishingwan.github.io/PRS-Tutorial/plink/.) The clumping cut-off of r^2^ was 0.1. The *p*-value threshold was 0.05 for the subjects in the training, validation, and test sets. The PRS of the subjects in the training set were then used to train a separate logistic regression classifier using the *glm* function in R (www.R-project.org) for each *p* value threshold. The validation data set was used to determine which of these thresholds produces the best classifier, which was then used to predict the test set.

### Deep learning

The deep learning prediction model was built using a Diet Network according to the procedure described by Romero et al.. (Romero et al., 2016) The official code can be found here: https://github.com/adri-romsor/DietNetworks.

### Feature selection

A knowledge graph containing contextual information mined from public databases such as e.g. dbSNP, ClinVar, OMIM, Reactome, STRING database proposes 100 SNPs and SNP-SNP interactions. The proposed SNPs and SNP-SNP interactions are evaluated in the training data by drop one model comparison procedure using the *glm* function in R (www.R-project.org). (Klinger et al.) A gradient descent algorithm directs the search across the graph based on whether proposed hypotheses are correlated with the disease status or not and corrects accordingly the SNP and SNP-SNP interactions list until all members of the list show strong correlation with the disease status.

### LASSO regression

LASSO regression models were computed by using the glmnet package (https://glmnet.stanford.edu/index.html for R (www.R-project.org) and its function cv.glmnet. (Friedman et al., 2010)

## Data Availability

Following publication of major outputs all anonymized data will be made available on reasonable request to the corresponding author providing this meets local ethical and research governance criteria.

## Acknowledgements

Data used in the preparation of this article were obtained from the Parkinson’s Progression Markers Initiative (PPMI) database (www.ppmi-info.org/data). For up-to-date information on the study, visit www.ppmi-info.org. PPMI – a public-private partnership – is funded by The Michael J. Fox Foundation for Parkinson’s Research and funding partners. List the full names of all of the PPMI funding partners can be found at www.ppmi-info.org/fundingpartners. The research work was supported by the Investitionsbank des Landes Brandenburg (ILB), the European Regional Development Fund (ERDF), and the European Social Fund+ (ESF+). We also thank the program digital solutions made in Brandenburg (digisolBB) for its continued support.

## References

Bateson, and William (1906). The progress of genetics since the rediscovery of Mendel’s papers. Progress. Rei Bot. 1, 368.

Butera, G., Mullappilly, N., Masetto, F., Palmieri, M., Scupoli, M. T., Pacchiana, R., et al. (2019). Regulation of Autophagy by Nuclear GAPDH and Its Aggregates in Cancer and Neurodegenerative Disorders. Int. J. Mol. Sci. 20. doi:10.3390/ijms20092062.

Chatterjee, N., Wheeler, B., Sampson, J., Hartge, P., Chanock, S. J., and Park, J.-H. (2013). Projecting the performance of risk prediction based on polygenic analyses of genome-wide association studies. Nat. Genet. 45, 400–5, 405e1–3.

Choi, S. W., Mak, T. S.-H., and O’Reilly, P. F. (2020). Tutorial: a guide to performing polygenic risk score analyses. Nat. Protoc. 15, 2759–2772.

Diogo, D., Tian, C., Franklin, C. S., Alanne-Kinnunen, M., March, M., Spencer, C. C. A., et al. (2018). Phenome-wide association studies across large population cohorts support drug target validation. Nat. Commun. 9, 4285.

Dudbridge, F. (2013). Power and predictive accuracy of polygenic risk scores. PLoS Genet. 9, e1003348.

Duncan, L., Shen, H., Gelaye, B., Meijsen, J., Ressler, K., Feldman, M., et al. (2019). Analysis of polygenic risk score usage and performance in diverse human populations. Nat. Commun. 10, 3328.

Evans, D. M., Visscher, P. M., and Wray, N. R. (2009). Harnessing the information contained within genome-wide association studies to improve individual prediction of complex disease risk. Hum. Mol. Genet. 18, 3525–3531.

Friedman, J., Hastie, T., and Tibshirani, R. (2010). Regularization Paths for Generalized Linear Models via Coordinate Descent. J. Stat. Softw. 33, 1–22.

Gola, D., Erdmann, J., Müller-Myhsok, B., Schunkert, H., and König, I. R. (2020). Polygenic risk scores outperform machine learning methods in predicting coronary artery disease status. Genet. Epidemiol. 44, 125–138.

International Schizophrenia Consortium, Purcell, S. M., Wray, N. R., Stone, J. L., Visscher, P. M., O’Donovan, M. C., et al. (2009). Common polygenic variation contributes to risk of schizophrenia and bipolar disorder. Nature 460, 748–752.

Jinn, S., Blauwendraat, C., Toolan, D., Gretzula, C. A., Drolet, R. E., Smith, S., et al. (2019). Functionalization of the TMEM175 p.M393T variant as a risk factor for Parkinson disease. Hum. Mol. Genet. 28, 3244–3254.

Jinn, S., Drolet, R. E., Cramer, P. E., Wong, A. H.-K., Toolan, D. M., Gretzula, C. A., et al. (2017). TMEM175 deficiency impairs lysosomal and mitochondrial function and increases α-synuclein aggregation. Proc. Natl. Acad. Sci. U. S. A. 114, 2389– 2394.

Klinger, J., Ravarani, C., Bannard, C., Lamparter, M., Schwinges, A., Cope, J., et al. Critically ill COVID-19 status associated trait genetics reveals CDK6 inhibitors as potential treatment. doi:10.21203/rs.3.rs-568366/v1.

Krohn, L., Öztürk, T. N., Vanderperre, B., Ouled Amar Bencheikh, B., Ruskey, J. A., Laurent, S. B., et al. (2020). Genetic, Structural, and Functional Evidence Link TMEM175 to Synucleinopathies. Ann. Neurol. 87, 139–153.

Lo, A., Chernoff, H., Zheng, T., and Lo, S.-H. (2015). Why significant variables aren’t automatically good predictors. Proc. Natl. Acad. Sci. U. S. A. 112, 13892–13897.

Lynch-Day, M. A., Mao, K., Wang, K., Zhao, M., and Klionsky, D. J. (2012). The role of autophagy in Parkinson’s disease. Cold Spring Harb. Perspect. Med. 2, a009357.

Manolio, T. A., Collins, F. S., Cox, N. J., Goldstein, D. B., Hindorff, L. A., Hunter, D. J., et al. (2009). Finding the missing heritability of complex diseases. Nature 461, 747–753.

Marees, A. T., de Kluiver, H., Stringer, S., Vorspan, F., Curis, E., Marie-Claire, C., et al. (2018). A tutorial on conducting genome-wide association studies: Quality control and statistical analysis. Int. J. Methods Psychiatr. Res. 27, e1608.

Marek, K., Chowdhury, S., Siderowf, A., Lasch, S., Coffey, C. S., Caspell-Garcia, C., et al. (2018). The Parkinson’s progression markers initiative (PPMI) - establishing a PD biomarker cohort. Ann Clin Transl Neurol 5, 1460–1477.

Marek, K., Jennings, D., Lasch, S., Siderowf, A., Tanner, C., Simuni, T., et al. (2011). The Parkinson Progression Marker Initiative (PPMI). Prog. Neurobiol. 95, 629– 635.

Nalls, M. A., Pankratz, N., Lill, C. M., Do, C. B., Hernandez, D. G., Saad, M., et al. (2014). Large-scale meta-analysis of genome-wide association data identifies six new risk loci for Parkinson’s disease. Nat. Genet. 46, 989–993.

Olanow, C. W., Schapira, A. H. V., LeWitt, P. A., Kieburtz, K., Sauer, D., Olivieri, G., et al. (2006). TCH346 as a neuroprotective drug in Parkinson’s disease: a double-blind, randomised, controlled trial. Lancet Neurol. 5, 1013–1020.

Purcell, S., Neale, B., Todd-Brown, K., Thomas, L., Ferreira, M. A. R., Bender, D., et al. (2007). PLINK: a tool set for whole-genome association and population-based linkage analyses. Am. J. Hum. Genet. 81, 559–575.

Reisberg, S., Iljasenko, T., Läll, K., Fischer, K., and Vilo, J. (2017). Comparing distributions of polygenic risk scores of type 2 diabetes and coronary heart disease within different populations. PLoS One 12, e0179238.

Romero, A., Carrier, P. L., Erraqabi, A., Sylvain, T., Auvolat, A., Dejoie, E., et al. (2016). Diet Networks: Thin Parameters for Fat Genomics. arXiv [cs.LG]. Available at: http://arxiv.org/abs/1611.09340.

Tibshirani, R. (1996). Regression shrinkage and selection via the lasso. J. R. Stat. Soc. 58, 267–288.

Wei, W.-H., Hemani, G., and Haley, C. S. (2014). Detecting epistasis in human complex traits. Nat. Rev. Genet. 15, 722–733.

Wray, N. R., Goddard, M. E., and Visscher, P. M. (2007). Prediction of individual genetic risk to disease from genome-wide association studies. Genome Res. 17, 1520–1528.

Yang, J., Benyamin, B., McEvoy, B. P., Gordon, S., Henders, A. K., Nyholt, D. R., et al. (2010). Common SNPs explain a large proportion of the heritability for human height. Nat. Genet. 42, 565–569.

Zhou, W., Nielsen, J. B., Fritsche, L. G., Dey, R., Gabrielsen, M. E., Wolford, B. N., et al. (2018). Efficiently controlling for case-control imbalance and sample relatedness in large-scale genetic association studies. Nat. Genet. 50, 1335–1341.

